# Cancer inpatient with COVID-19: a report from the Brazilian National Cancer Institute

**DOI:** 10.1101/2020.06.27.20141499

**Authors:** Andreia C. de Melo, Luiz C.S. Thuler, Jesse L. da Silva, Lucas Z. de Albuquerque, Ana C. Pecego, Luciana de O.R. Rodrigues, Magda S. da Conceição, Marianne M. Garrido, Gelcio L. Quintella Mendes, Ana Cristina P. Mendes Pereira, Marcelo A. Soares, João P.B. Viola, on behalf of the INCA COVID-19 Task Force

## Abstract

Brazil has been recording a frightening exponential curve of confirmed cases of SARS-CoV-2 infection. Cancer patients with COVID-19 are likely to have a greater risk of complications and death. A retrospective search in the electronic medical records of cancer inpatients admitted to the Brazilian National Cancer Institute from April 30, 2020 to May 26, 2020 granted identification of 181 patients with COVID-19 confirmed by RT-PCR method. The mean age was 55.3 years (SD ±21.1). The most prevalent solid tumors were breast (40 [22.1%]), gastrointestinal (24 [13.3%]), and gynecological (22 [12.2%]). Among hematological malignancies, lymphoma (20 [11%]) and leukemia (10 [5.5%]) predominated. The most common complications were respiratory failure (70 [38.7%]), septic shock (40 [22.1%]) and acute kidney injury (33 [18.2%]). A total of 60 (33.1%) patients died due to COVID-19 complications. By multivariate analysis, cases with admission due to symptoms of COVID-19 (p = 0.027) and with two or more metastatic sites (p <0.001) showed a higher risk of COVID-19-specific death. This is the first study in a cohort of Brazilian cancer patients with COVID-19. The rates of complications and COVID-19-specific death were significantly high. Our data prompts urgent and effective public policies for this group of especially vulnerable patients.

**Statement of Significance:** COVID-19-specific mortality in cancer inpatients is markedly higher than in the general population and the cases with advanced cancer are particularly in a more vulnerable group. Adaptations of cancer management guidelines and more intensive preventive measures should be a priority for this group of patients.

## Introduction

The novel coronavirus, named severe acute respiratory syndrome coronavirus-2 (SARS-CoV-2) that causes the coronavirus disease 2019 (COVID-19)^1^, was first detected in Wuhan, the provincial capital of Hubei, China, in December 2019. SARS-CoV-2 has rapidly spread to many other countries worldwide becoming an unprecedented astounding and devastating pandemic in a short period of time.

Following an exponential upward trend, the increasing number of cases and death toll remain to concern the scientific community around the globe. Currently, more than 8.99 million cases are confirmed worldwide, with more than 469,500 deaths^2^. The first case of COVID-19 in Brazil was detected on February 26, 2020. Standing out worldwide for having one of the steepest epidemiological curves, the country has already reached the second place in incidence with almost 1.08 million cases and second place in mortality with more than 50,500 deaths so far^3^. As the most populous areas, the states of São Paulo and Rio de Janeiro have predictably concentrated the highest numbers of cases and deaths. Thus far, Brazil has been considered the new epicenter of the global pandemic^4^.

In general, the vast majority of COVID-19 patients develop mild symptoms or remain asymptomatic over the course of the disease. An intermediate group of patients have moderate symptoms requiring hospitalization and some noninvasive intervention. A few number of patients have a more severe course of the disease with desaturation, dyspnea, septic shock, and/or multiple organ dysfunction leading to life-threatening consequences or death^5^.

Patients with cancer are more likely to have severe complications and even death when affected by COVID-19^6–8^, mainly due to the effects of the immunosuppressive anticancer treatments, frequent use of corticosteroids, advanced age, comorbidities and pulmonary involvement (primary tumors or secondary lung metastases). Particularly in low- and middle-income countries, COVID-19 has brought a heavy burdening to the public health systems and induced new planning and adjustments in the clinical approach to cancer patients^9^.

Based on a few series previously published around the world, data evaluating the impact of COVID-19 outbreak in management and survival of patients with cancer are still very scarce, incomplete, with heterogeneous outcomes and descriptions^10–15^. Brazilian data in this specific field are still unknown due to the lack of publications.

The aim of this report was to describe the demographics, clinical characteristics and laboratory abnormalities of cancer inpatients with COVID-19 admitted to the hospital ward of the Brazilian National Cancer Institute (INCA), exploring factors associated with death.

## Methods

### Study design and participants

This retrospective cohort was performed through a search on electronic medical records and compiled data of cancer inpatients admitted to INCA with laboratory-confirmed SARS-CoV-2 infection between April 30, 2020 and May 26, 2020. The hospital admission occurred for various medical reasons, including COVID-19 symptoms or any other clinical condition (for those cases with onset of symptoms throughout hospitalization or cancer inpatients who had contact to other COVID-19 cases). Outpatients tested positive for SARS-CoV-2 infection and patients with only non-invasive cancer (or pre-malignant conditions) were not the object of this study.

COVID-19 was diagnosed on the basis of the WHO interim guidance^16^, in which confirmation was defined as a positive result on real-time reverse transcriptase polymerase chain reaction (RT-PCR) assay of nasal- and oropharyngeal swab specimens using the U.S. Centers for Disease Control and Prevention (CDC) reagents and protocol^17^. Specimens were collected right after the hospital admission from those patients with COVID-19 symptoms and immediately after clinical suspicion from those admitted to hospital for diverse reasons unrelated to COVID-19.

The study was approved by the National Commission of Ethics in Research (CONEP) and conducted in accordance with the Good Clinical Practice guidelines. Written informed consent was waived due to the retrospective design and the emergency feature of the research. Only anonymized data was analyzed.

### Data collection and outcomes

The demographic and clinical characteristics, including tumor site, histological subtype, staging, site of metastases, cancer treatment within the last 60 days, the presence of comorbidities, COVID-19-related clinical signs and symptoms, and laboratory tests at diagnosis and throughout hospitalization were obtained from the electronic medical records. COVID-19-specific clinical treatments were also collected. The variables analyzed in order to feature disease severity were admission to the intensive care unit (ICU), mechanical ventilation, renal failure, hemodialysis, septic shock, and death. Patients transferred out from INCA to another hospital were censored on the date of transfer. Patients who had not been discharged from hospital were censored in the date of the last follow-up on May 31, 2020.

### Statistical analysis

The statistical software package SPSS, version 21.0 (São Paulo, Brazil) was used for the analyses. All continuous variables were evaluated by the Kolmogorov-Smirnov test of normality. Categorical variables were shown in percentages or absolute values. The study endpoint was COVID-19-related mortality. Time of follow-up was calculated from the date of swab collection to hospital discharge, death, or censorship of patients who were transferred or still hospitalized at the end of the study.

Risk factors for death were assessed using logistic regression. Crude and adjusted odds ratios (OR) were calculated. Variables with a p-value <0.20 at the univariate analysis were included in the multivariate model by stepwise forward selection with the entry order based on their level of significance. All p-values <0.05 were considered statistically significant.

## Results

A total of 181 patients had the diagnosis of COVID-19 confirmed at INCA and were considered eligible for this study (Figure 1). The median follow-up for the general population was 5 days (interquartile range, IQR 2-10.3).

**Figure 1.**
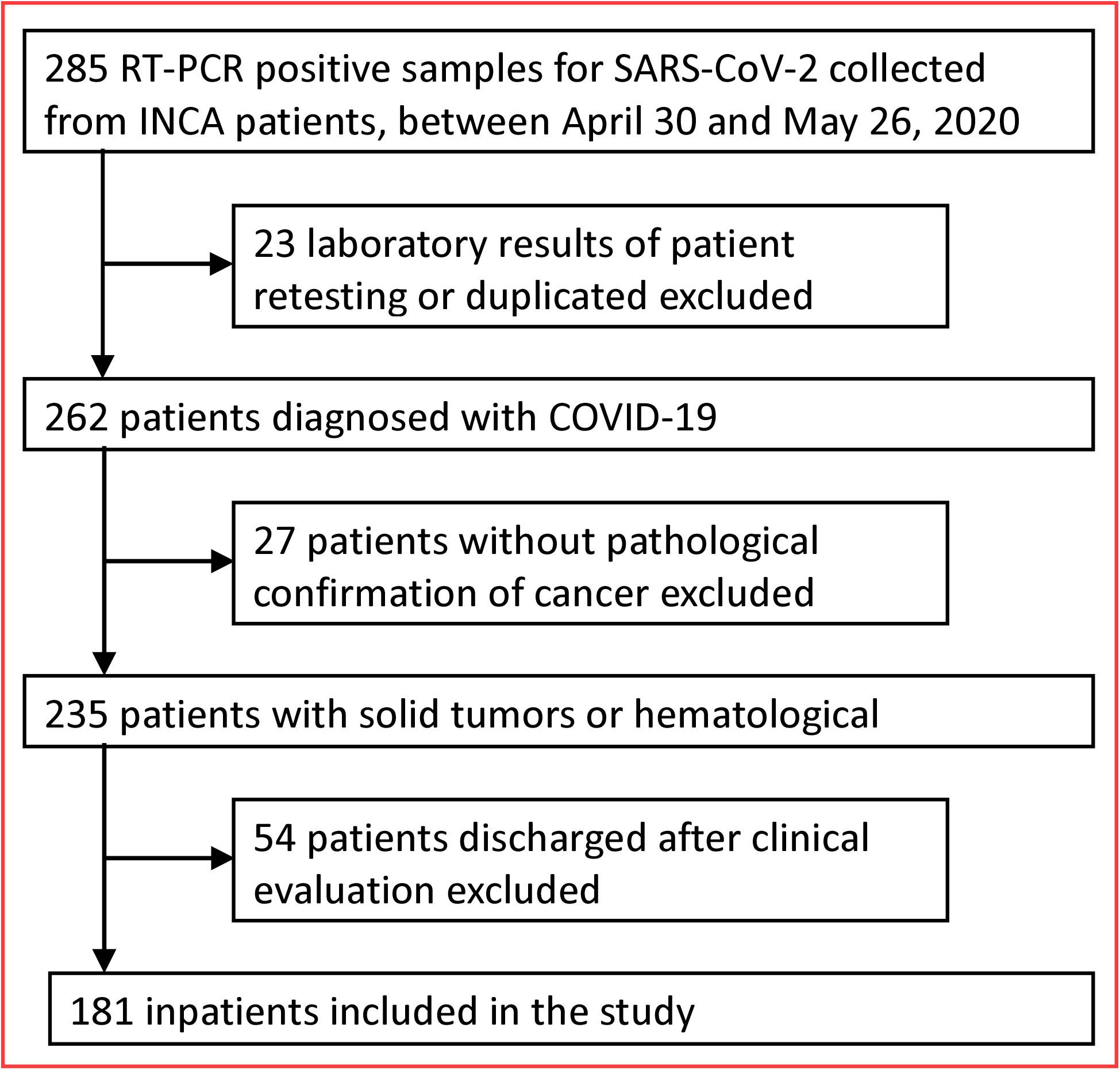
Flow chart of study patient inclusion

The clinical and demographic characteristics are described in detail in Table 1. The mean age was 55.3 years (standard deviation, SD ±21.1) and 92 (50.8%) patients aged 60 or older. Female gender was more prevalent (110 [60.8%]) and 40 patients (22.1%) were former or current smokers. Comorbidities were found in 110 (60.8%) cases, of which the most common were hypertension (77 [42.5%]) and diabetes (31 [17.1%]), and 21 (11.6%) patients had three or more comorbidities. Long-term use of corticosteroids was seen in 21 (11.6%) cases.

**Table 1.** Baseline demographic and clinical characteristics of the patients

|  | **N (%)** |  |
| --- | --- | --- |
| **Variables** |  |  |
| **Age, years** |  |  |
| Mean (±SD) | 55.3 (±21.1) | |
| Range | 1.8 - 88.0 |  |
| < 60 | 89 | (49.2) |
| 60 - 74 | 67 | (37.0) |
| ≥ 75 | 25 | (13.8) |
| **Sex** |  |  |
| Female | 110 | (60.8) |
| Male | 71 | (39.2) |
| **Smoking status** |  |  |
| Never | 69 | (38.1) |
| Current | 18 | (9.9) |
| Former | 22 | (12.2) |
| Missing | 72 | (39.8) |
| **Comorbidities** |  |  |
| No | 46 | (25.4) |
| Yes | 110 | (60.8) |
| Missing | 25 | (13.8) |
| **Main Comorbidities** |  |  |
| Hypertension | 77 | (42.5) |
| Diabetes | 31 | (17.1) |
| Chronic renal failure | 10 | (5.5) |
| COPD/asthma | 7 | (3.9) |
| Other* | 21 | (11.6) |
| **Number of comorbidities** |  |  |
| 0 | 46 | (25.4) |
| 1 | 61 | (33.7) |
| 2 | 28 | (15.5) |
| ≥ 3 | 21 | (11.6) |
| Missing | 25 | (13.8) |
| **Long-term systemic corticosteroid use** |  |  |
| No | 143 | (79.0) |
| Yes | 21 | (11.6) |
| Missing | 17 | (9.4) |
| **Total** | **181** |  |

\*Congestive heart failure, arrhythmia, ischemic heart disease, cerebrovascular disease, morbid obesity (BMI > 40 Kg/m<sup>2</sup>), HIV infection. SD=standard deviation. COPD=chronic obstructive pulmonary disease.

The baseline oncological characteristics of the study patients are displayed in Table 2. The median time from cancer diagnosis to COVID-19 was 1.5 years (IQR 0.3-4.1). The most prevalent solid tumors were breast (40 [22.1%]), gastrointestinal (24 [13.3%]), gynecological (22 [12.2%]) and urological (17 [9.4%]). Lung cancer patients made up only 7 (3.9%) cases. Among hematological malignancies, lymphoma (20 [11%]) and leukemia (10 [5.5%]) predominated. Patients with metastatic disease accounted for almost half of the cases (90 [49.7%]) and 32 (17.7%) patients had lung metastases. More than a third of the cases (63 [34.8%]) had recently undergone cytotoxic chemotherapy within the last 60 days before the COVID-19 diagnosis, 32 (17.7%) were in best supportive care and 27 (14.9%), in post-treatment clinical surveillance. Only nine patients (5%) were receiving targeted therapy or immunotherapy with checkpoint inhibitors as the current treatment line.

**Table 2.**
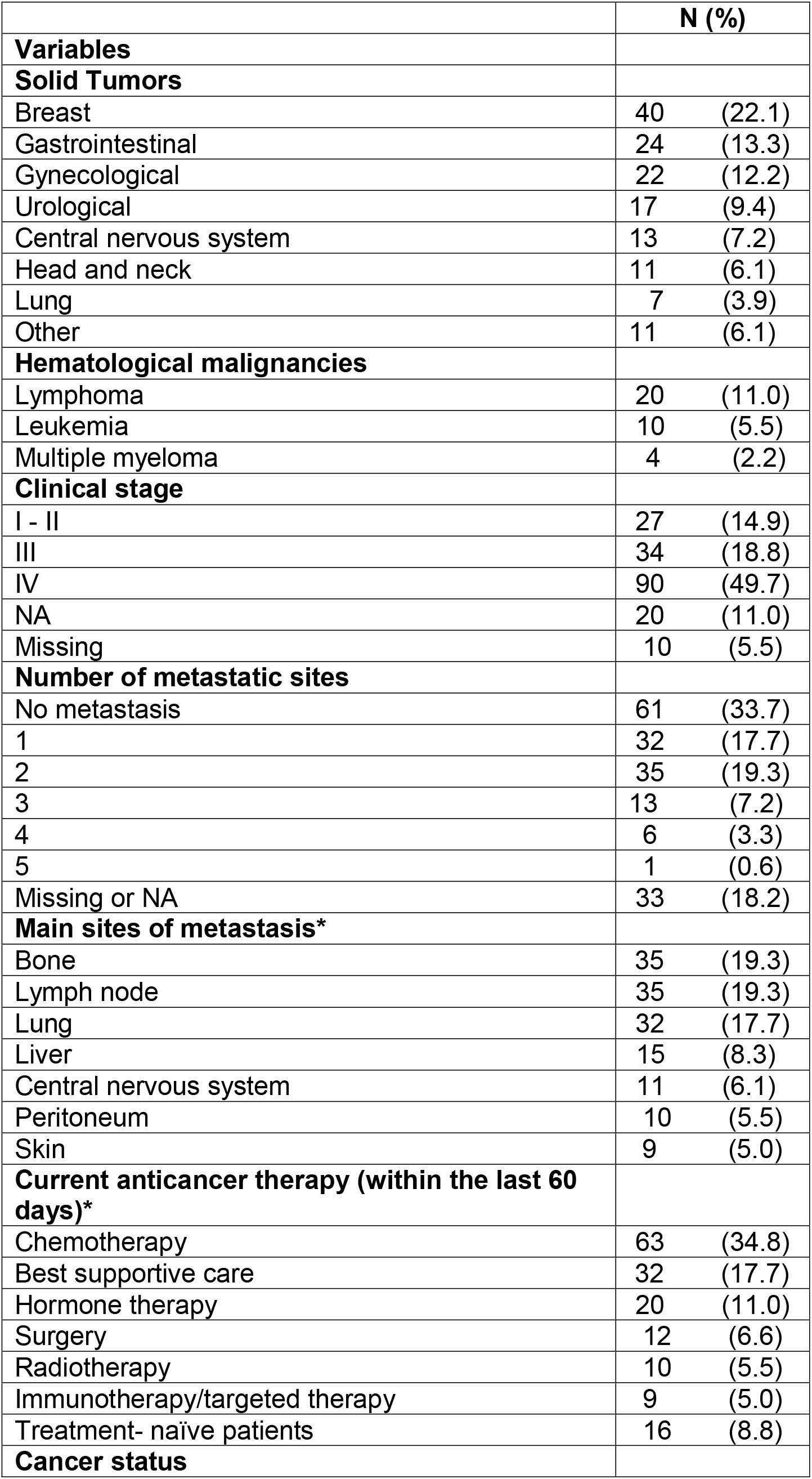
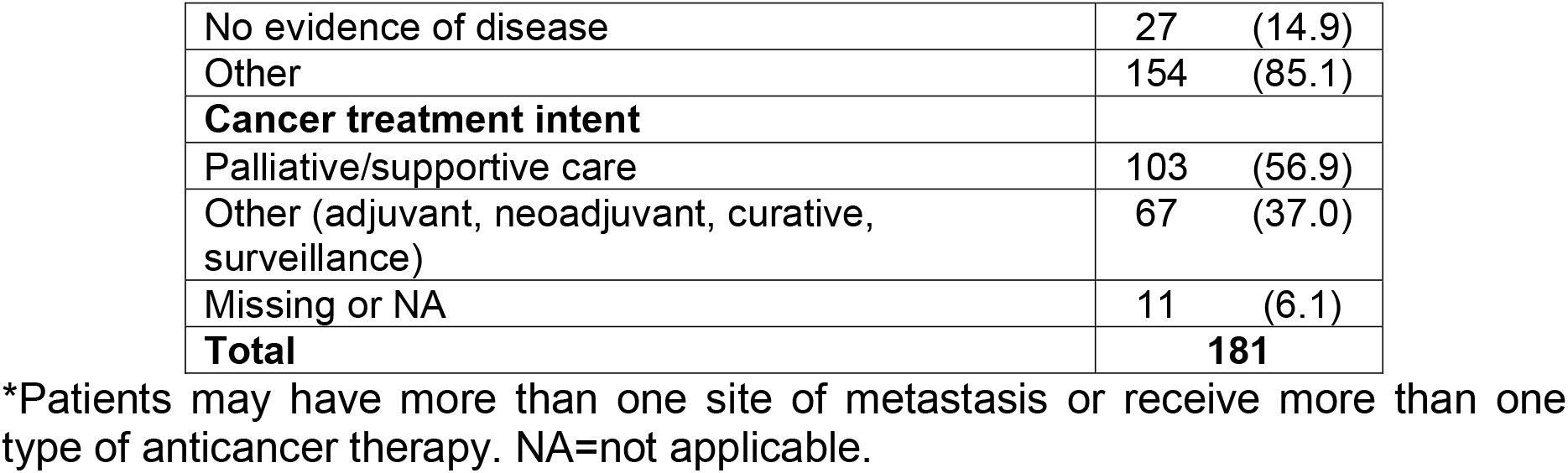
Cancer characteristics of the study patients

Information regarding the conditions of hospital admission and clinical evolution of patients are summarized in Table 3. More than half of the patients (98 [54.1%]) were admitted due to clinical worsening related to COVID-19 and 151 (83.4%) were symptomatic at the time of diagnostic confirmation. The most frequent symptoms were dyspnea (94 [51.9%]), cough (87 [48.1%]) and fever (66 [36.5%]). Admission to the ICU occurred in 32 (17.7%) cases, 130 (71.8%) patients required supplemental oxygen, and 35 (19.3%) cases progressed unfavorably with the need for mechanical ventilation. Ten patients (5.5%) were transferred out from INCA during the course of COVID-19. Figure 2A shows, from the day of swab collection, the timeline of events during hospital stay for COVID-19 patients and highlights that some patients had a rapidly deterioration of their clinical course once infected with SARS-CoV-2.

**Table 3.**
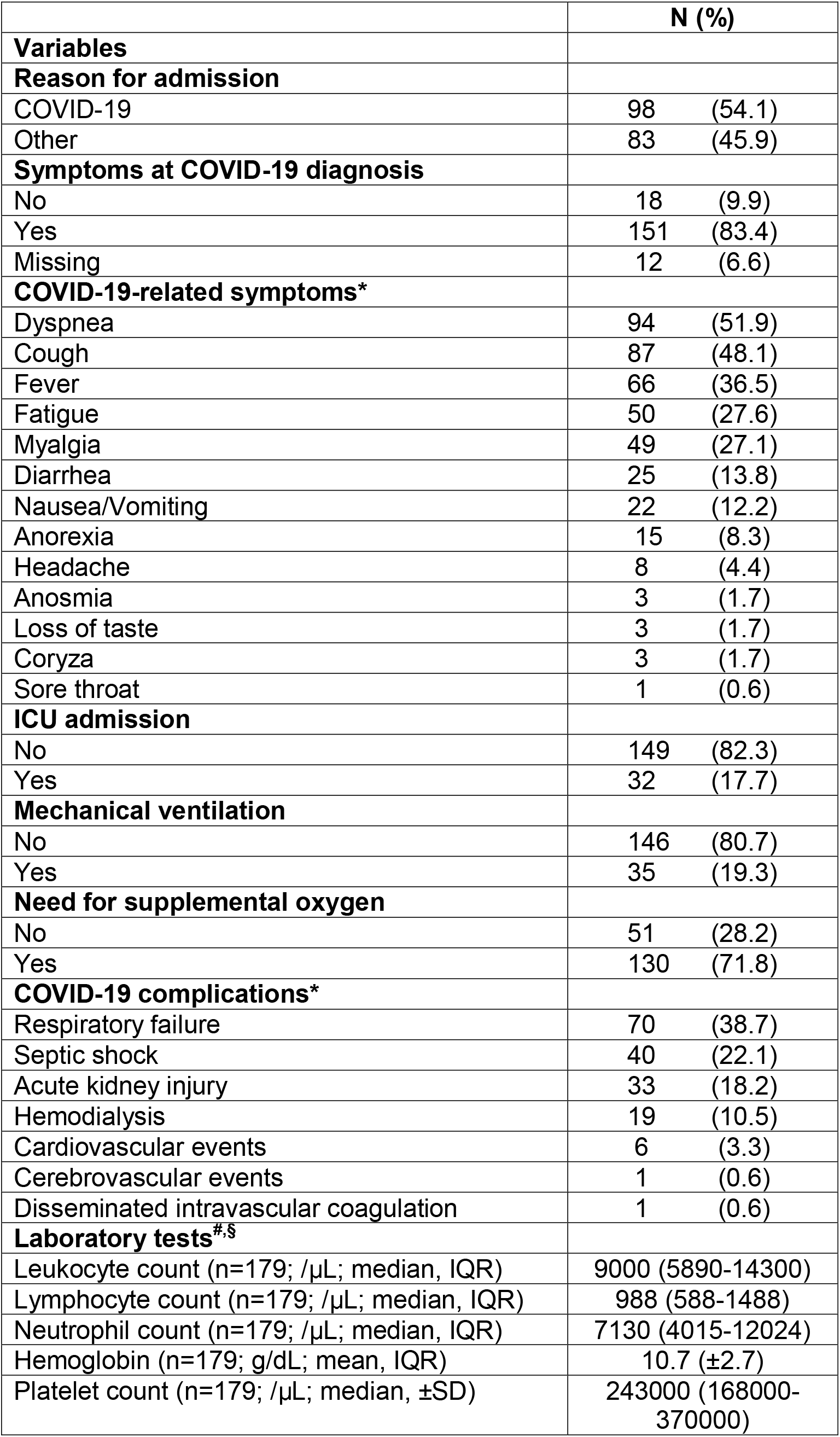
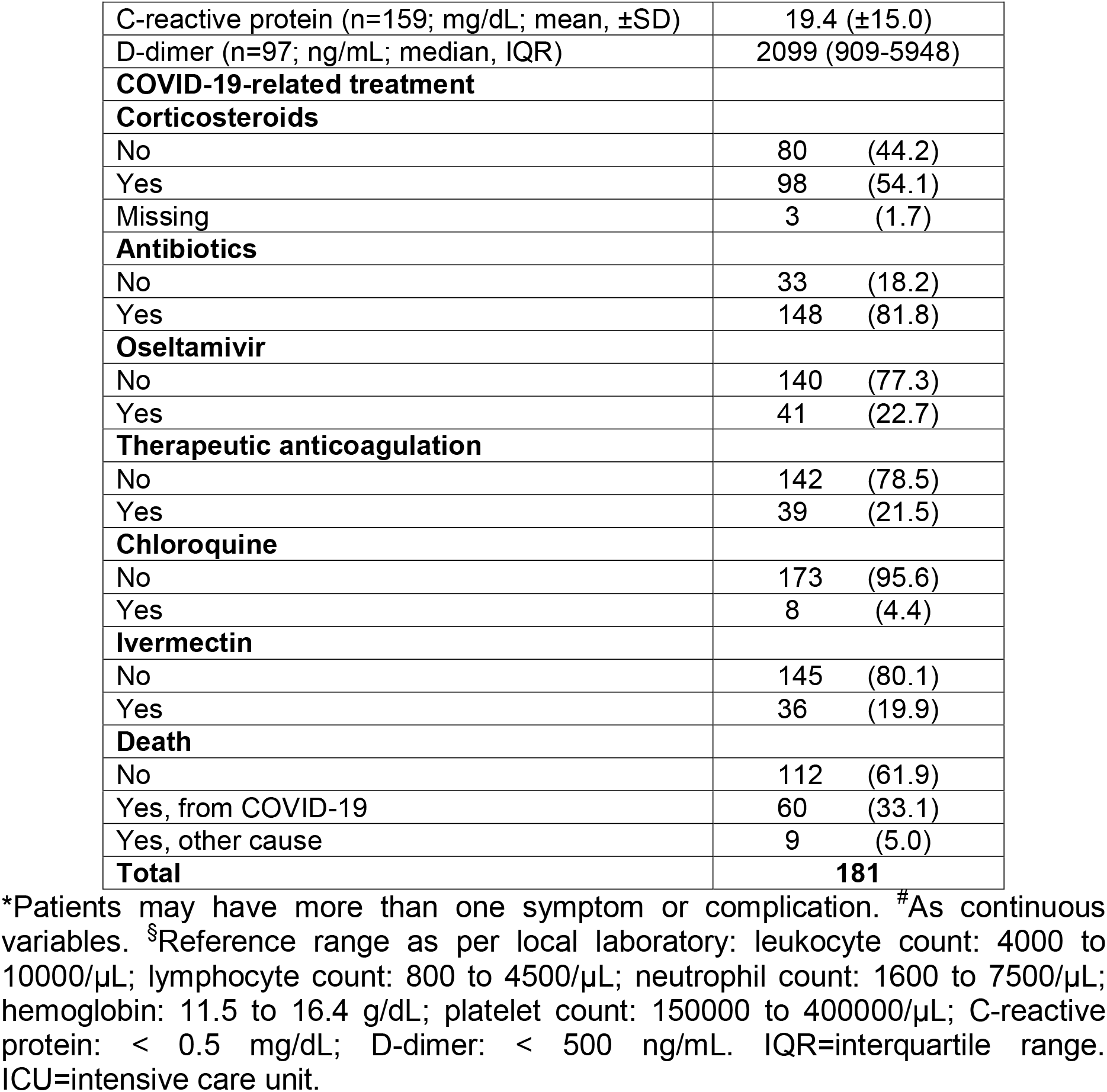
Characteristics of the patients’ current hospital admission

|  | **N (%)** |  |
| --- | --- | --- |
| **Variables** |  |  |
| **Reason for admission** |  |  |
| COVID-19 | 98 | (54.1) |
| Other | 83 | (45.9) |
| **Symptoms at COVID-19 diagnosis** |  |  |
| No | 18 | (9.9) |
| Yes | 151 | (83.4) |
| Missing | 12 | (6.6) |
| **COVID-19-related symptoms\*** |  |  |
| Dyspnea | 94 | (51.9) |
| Cough | 87 | (48.1) |
| Fever | 66 | (36.5) |
| Fatigue | 50 | (27.6) |
| Myalgia | 49 | (27.1) |
| Diarrhea | 25 | (13.8) |
| Nausea/Vomiting | 22 | (12.2) |
| Anorexia | 15 | (8.3) |
| Headache | 8 | (4.4) |
| Anosmia | 3 | (1.7) |
| Loss of taste | 3 | (1.7) |
| Coryza | 3 | (1.7) |
| Sore throat | 1 | (0.6) |
| **ICU admission** |  |  |
| No | 149 | (82.3) |
| Yes | 32 | (17.7) |
| **Mechanical ventilation** |  |  |
| No | 146 | (80.7) |
| Yes | 35 | (19.3) |
| **Need for supplemental oxygen** |  |  |
| No | 51 | (28.2) |
| Yes | 130 | (71.8) |
| **COVID-19 complications\*** |  |  |
| Respiratory failure | 70 | (38.7) |
| Septic shock | 40 | (22.1) |
| Acute kidney injury | 33 | (18.2) |
| Hemodialysis | 19 | (10.5) |
| Cardiovascular events | 6 | (3.3) |
| Cerebrovascular events | 1 | (0.6) |
| Disseminated intravascular coagulation | 1 | (0.6) |
| **Laboratory tests<sup>#,§</sup>** |  |  |
| Leukocyte count (n=179; /μL; median, IQR) | 9000 (5890-14300) | |
| Lymphocyte count (n=179; /μL; median, IQR) | 988 (588-1488) | |
| Neutrophil count (n=179; /μL; median, IQR) | 7130 (4015-12024) | |
| Hemoglobin (n=179; g/dL; mean, IQR) | 10.7 (±2.7) | |
| Platelet count (n=179; /μL; median, ±SD) | 243000 (168000-370000) | |

|  |  |
| --- | --- |
| C-reactive protein (n=159; mg/dL; mean, ±SD) | 19.4 (±15.0) |
| D-dimer (n=97; ng/mL; median, IQR) | 2099 (909-5948) |
| **COVID-19-related treatment** |  |
| **Corticosteroids** |  |
| No | 80 (44.2) |
| Yes | 98 (54.1) |
| Missing | 3 (1.7) |
| **Antibiotics** |  |
| No | 33 (18.2) |
| Yes | 148 (81.8) |
| **Oseltamivir** |  |
| No | 140 (77.3) |
| Yes | 41 (22.7) |
| **Therapeutic anticoagulation** |  |
| No | 142 (78.5) |
| Yes | 39 (21.5) |
| **Chloroquine** |  |
| No | 173 (95.6) |
| Yes | 8 (4.4) |
| **Ivermectin** |  |
| No | 145 (80.1) |
| Yes | 36 (19.9) |
| **Death** |  |
| No | 112 (61.9) |
| Yes, from COVID-19 | 60 (33.1) |
| Yes, other cause | 9 (5.0) |
| **Total** | **181** |

\*Patients may have more than one symptom or complication. <sup>#</sup>As continuous variables. <sup>s</sup>Reference range as per local laboratory: leukocyte count: 4000 to 10000/ μL; lymphocyte count: 800 to 4500/ μL; neutrophil count: 1600 to 7500/ μL; hemoglobin: 11.5 to 16.4 g/dL; platelet count: 150000 to 400000/ μL; C-reactive protein: < 0.5 mg/dL; D-dimer: < 500 ng/mL. IQR=interquartile range. ICU=intensive care unit.

**Figure 2.**
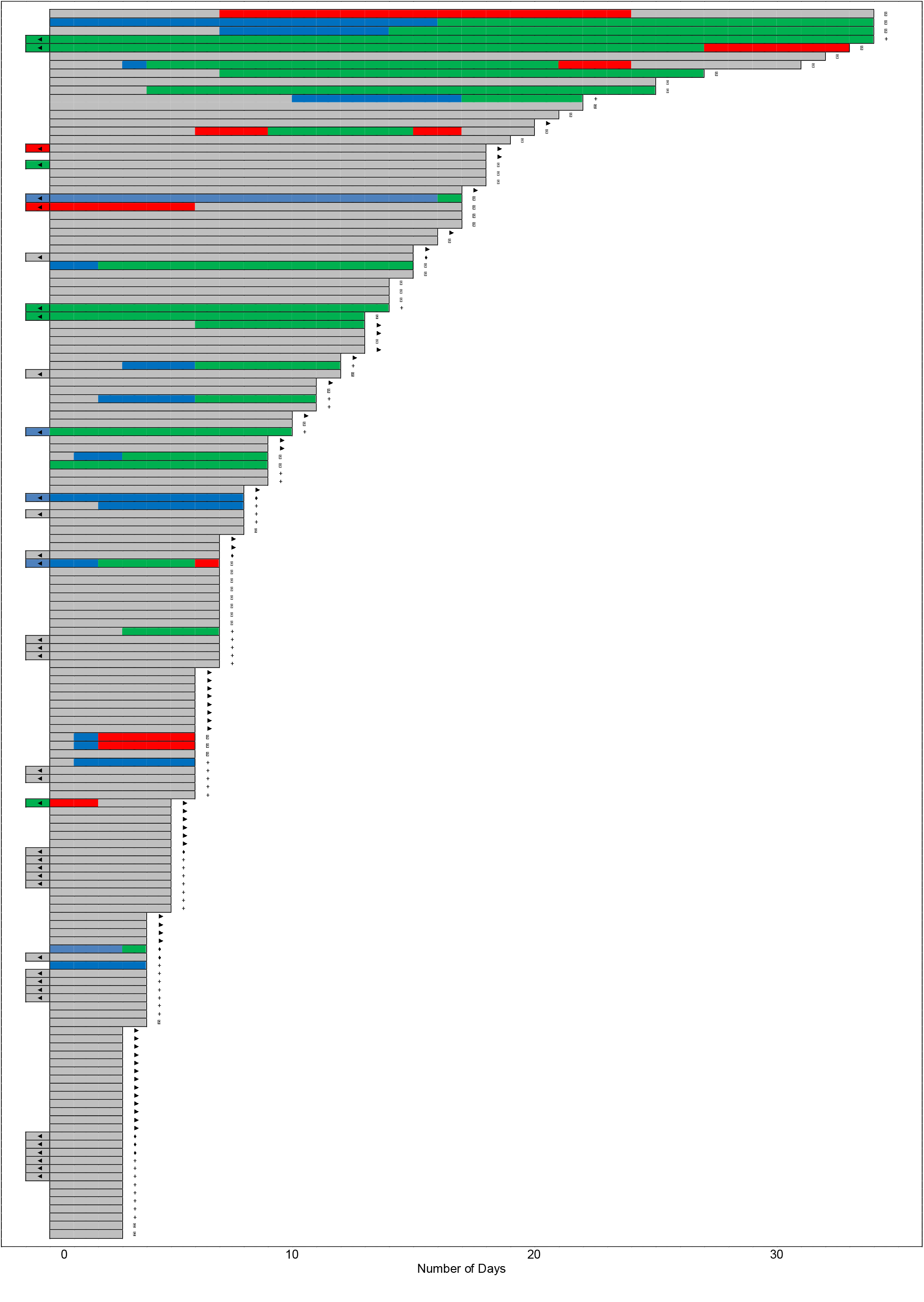
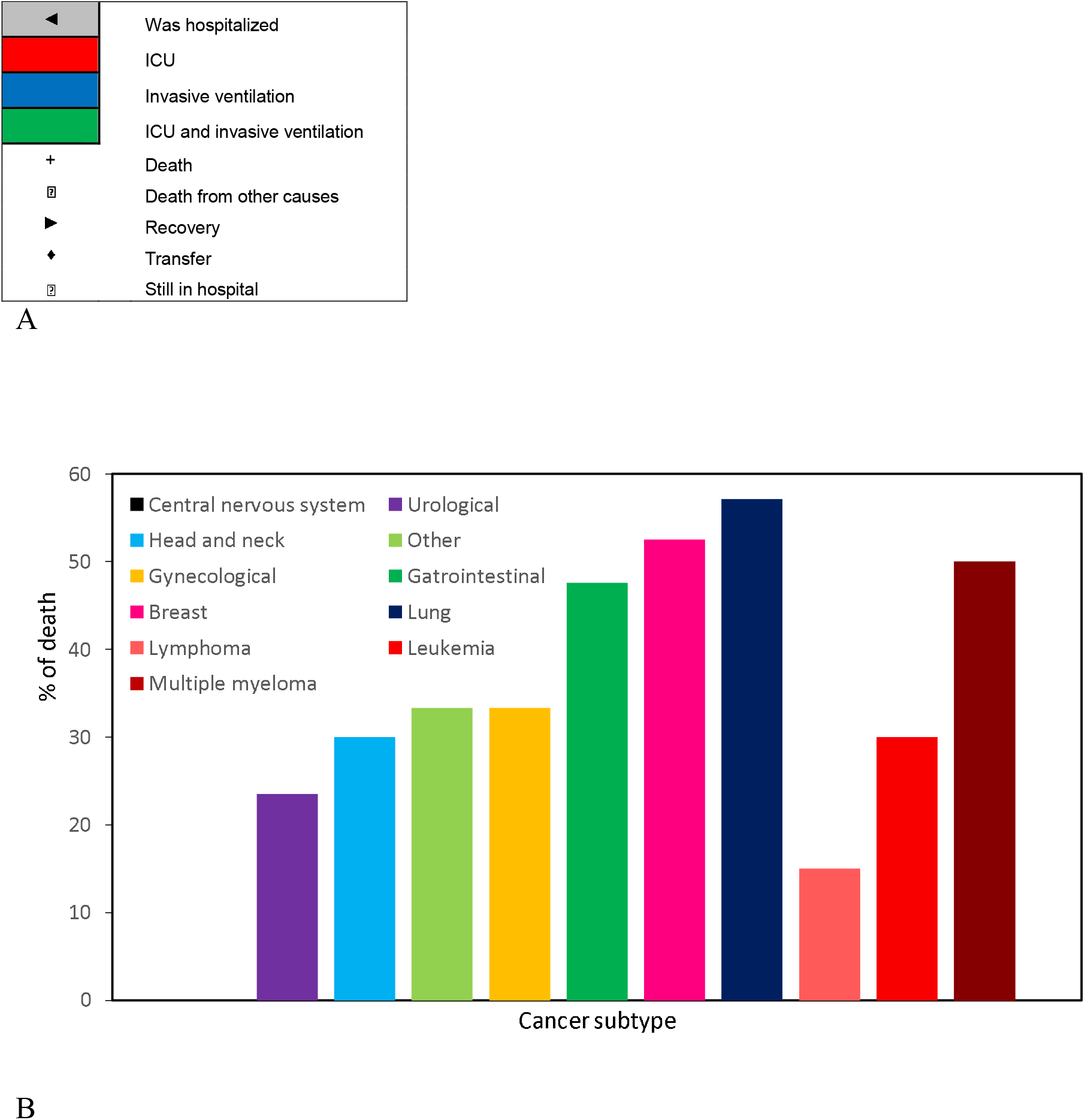
A: timeline of events during hospital stay for COVID-19 patients (from the day of swab collection). For visualization purposes, patients with short follow-up were excluded. B: percentage of death according to cancer subtype

The most common complications during hospitalization were respiratory failure (70 [38.7%]), septic shock (40 [22.1%]) and acute kidney injury (33 [18.2%]), with 19 (10.5%) patients requiring hemodialysis support. As for laboratory results, the median absolute lymphocytes count was 988/μL (IQR 588-1488), the mean C-reactive protein levels were 19.4 mg/dL (SD ±15.0) and median D-dimer levels were 2099 ng/mL (IQR 909-5948).

Under the clinical diagnosis of a severe acute respiratory syndrome, where influenza infection was considered one of the causative hypotheses and before the final RT-PCR result was confirmed as SARS-CoV-2 infection, oseltamivir was administered to 41 (22.7%) cases. During the course of COVID-19, more than half of the patients (98 [54.1%]) received corticosteroids, 148 (81.8%) were treated with antibiotics (including those against bacterial coinfections), and therapeutic anticoagulation was prescribed to 39 (21.5%) patients. Only eight (4.4%) patients received chloroquine and 36 (19.9%), ivermectin.

At the time of analysis, a total of 60 patients (33.1%) had died due to COVID-19. For solid tumors, the COVID-19-specific mortality rate was 37.7% (52/138) and for hematological malignancies (leukemia, lymphoma and multiple myeloma) was 23.5% (8/34). Four out of seven (57.1%) patients with lung cancer died from COVID-19, as well 52.5% (21/40) of breast cancer patients (Figure 2B).

As shown in Table 4, mortality related to COVID-19 was significantly associated to older age (p <0.001 for patients between 60 to 74 years and p = 0.002 for patients aged 75 years or older), metastatic cancer (p <0.001), two or more sites of metastases (p <0.001), the presence of lung (p <0.001) or bone metastases (p = 0.001), palliative or best supportive care intent (p <0.001), higher C-reactive protein levels (p = 0.002), admission due to COVID-19 (p = 0.009), and antibiotics use (p = 0.02). Isolated or combined comorbidities and elevated D-dimer levels did not demonstrate increased risk of dying from COVID-19.

**Table 4.**
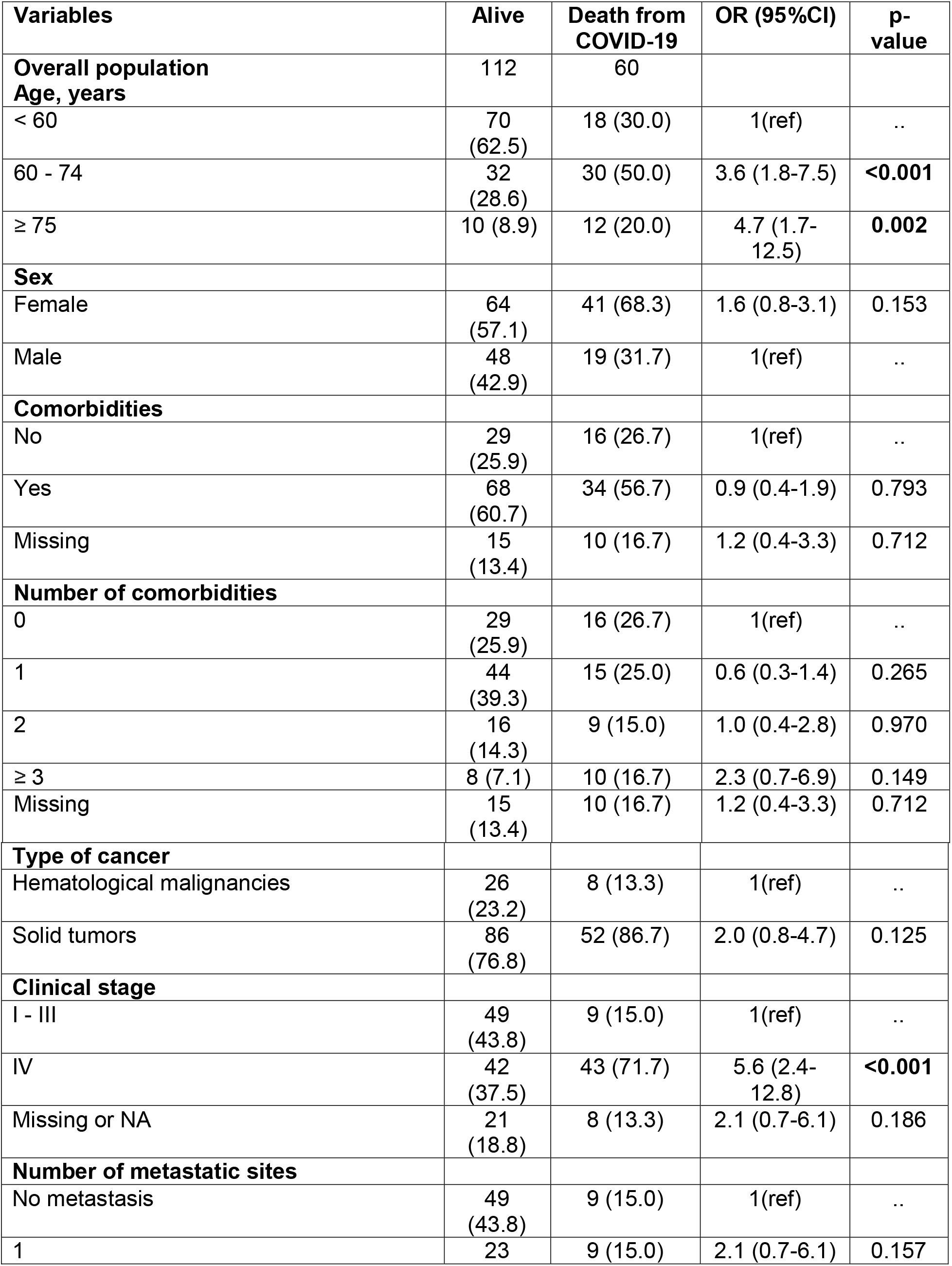
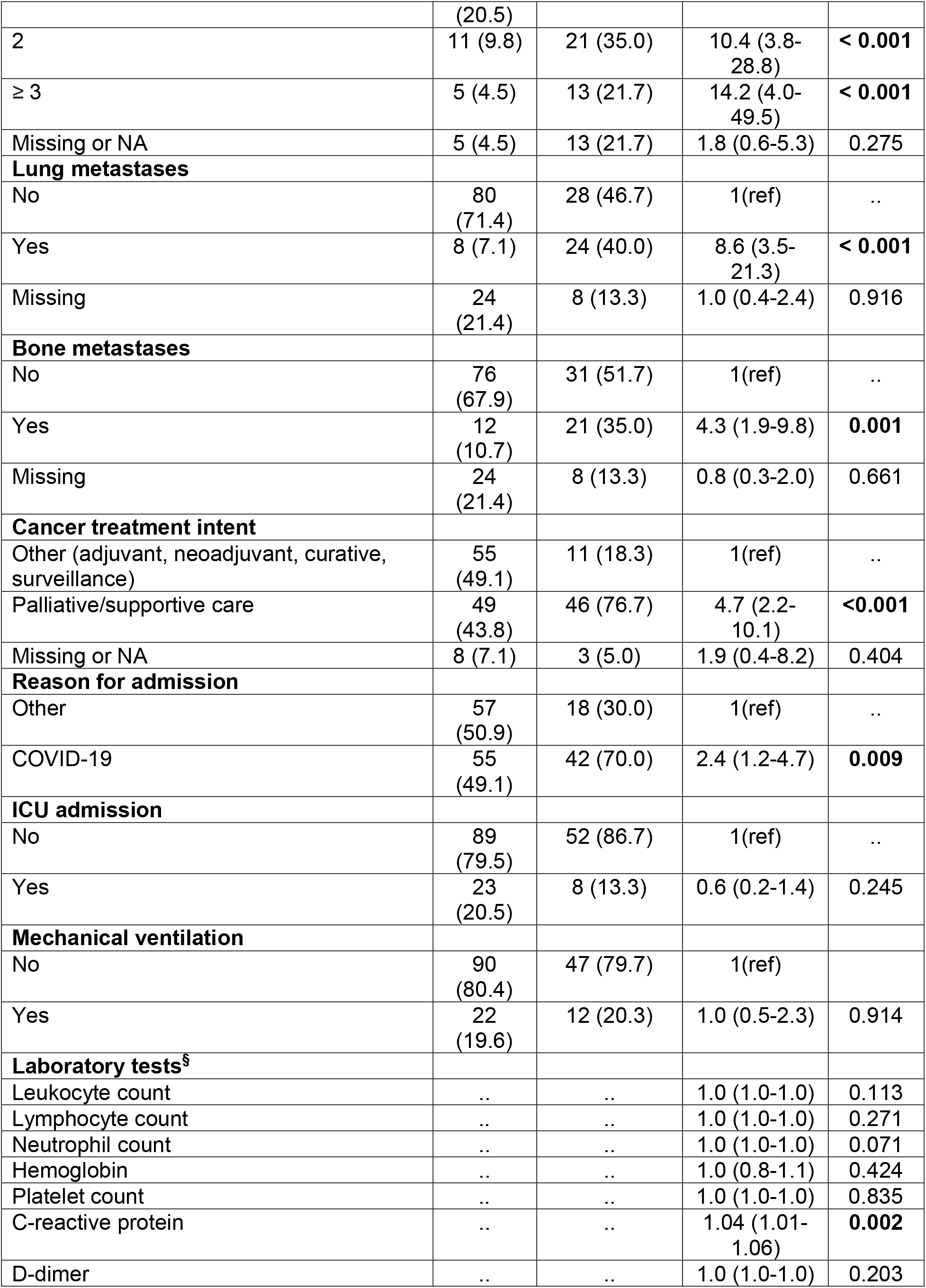
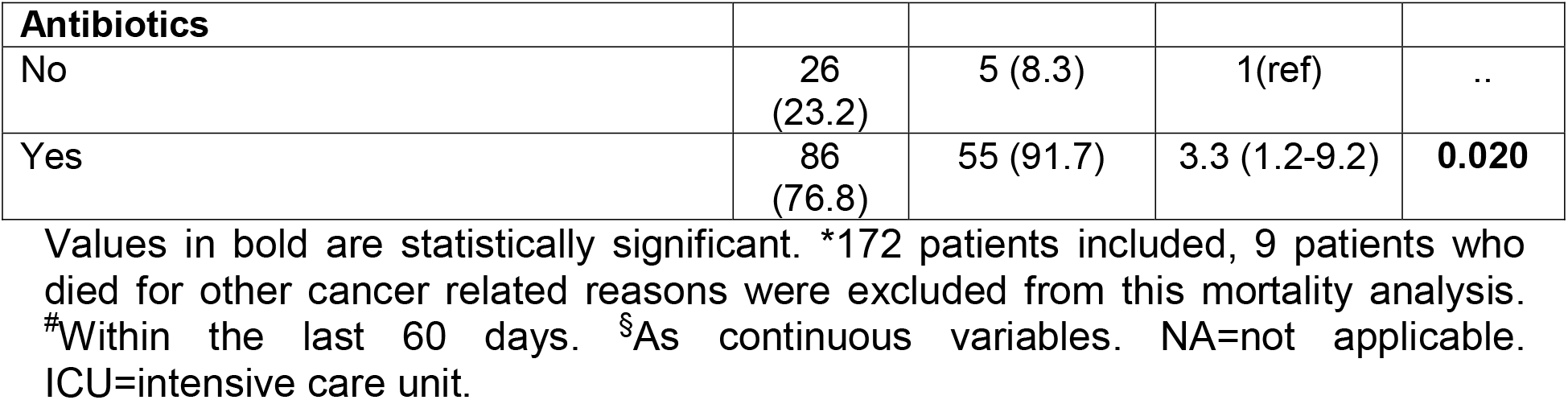
Variables associated to the risk of death from COVID-19*

Different modalities of cancer therapy, including systemic agents (chemotherapy, hormone therapy, targeted therapy, immunotherapy), surgical procedures or radiotherapy within 60 days before COVID-19 were not associated with mortality. Also, specific therapies during the COVID-19 course, such as oseltamivir, therapeutic anticoagulation, corticosteroids, ivermectin and chloroquine did not influence the risk of death (data not shown).

According to the multivariate analysis patients admitted due to COVID-19 symptoms (p = 0.027) and with two or more metastatic sites (p <0.001) showed a significantly higher risk of COVID-19-specific death (Table 5).

**Table 5.**
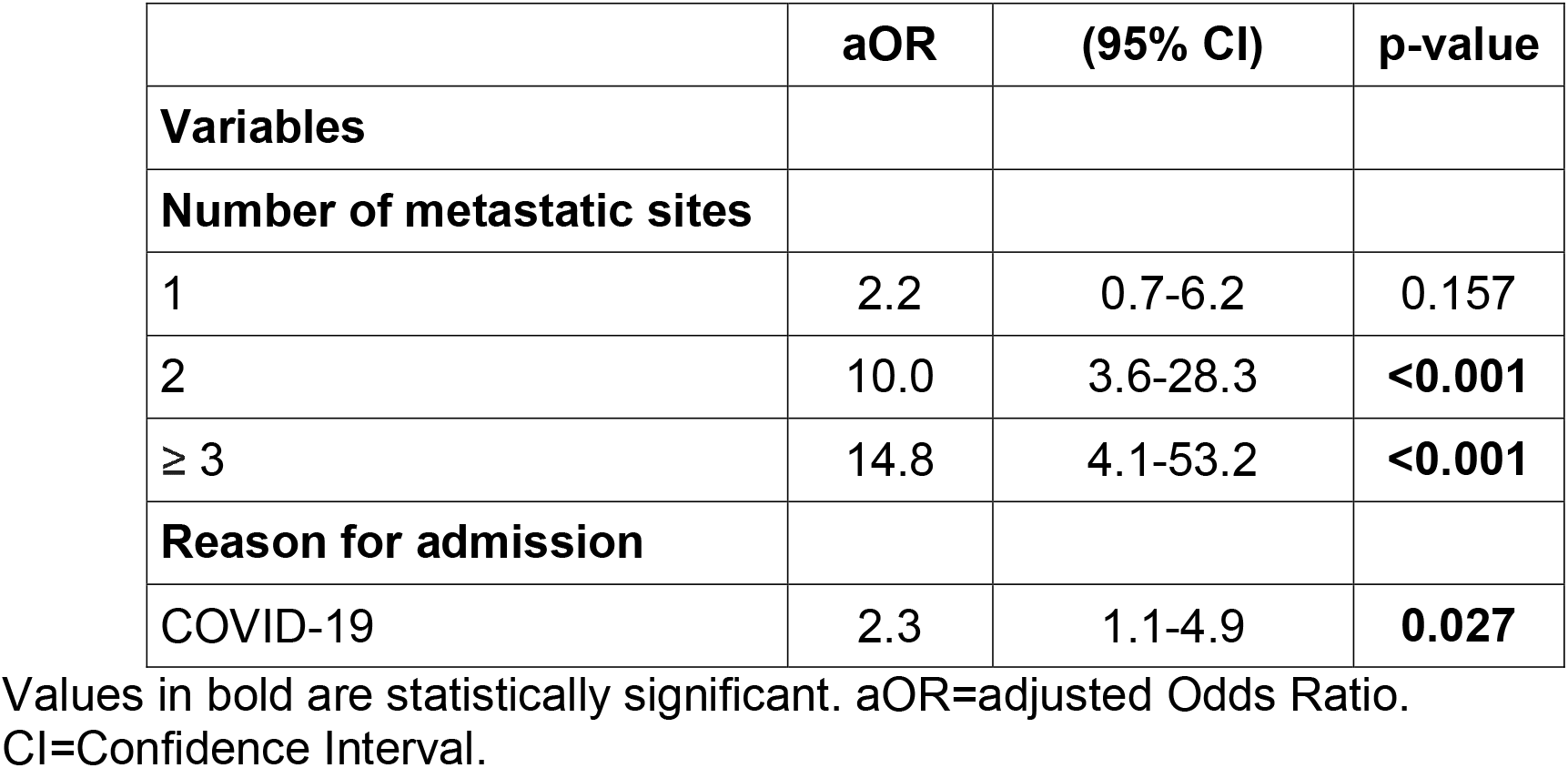
Independent variables associated with risk of death from COVID-19

|  | **aOR** | **(95% CI)** | **p-value** |
| --- | --- | --- | --- |
| **Variables** |  |  |  |
| **Number of metastatic sites** |  |  |  |
| 1 | 2.2 | 0.7-6.2 | 0.157 |
| 2 | 10.0 | 3.6-28.3 | **<0.001** |
| ≥ 3 | 14.8 | 4.1-53.2 | **<0.001** |
| **Reason for admission** |  |  |  |
| COVID-19 | 2.3 | 1.1-4.9 | **0.027** |

Values in bold are statistically significant. aOR=adjusted Odds Ratio. CI=Confidence Interval.

## Discussion

The findings of this cohort highlight, in detail, several significant aspects of the COVID-19 course in patients already diagnosed with cancer. Although the emergency period for case selection was considerably short, due to the large number of cancer patients admitted to INCA with COVID-19, 181 patients were successfully included for analysis. Women had greater representation, more than half of the patients aged 60 years or older and almost a quarter of the patients had also reported smoking. Patients with two or more comorbidities accounted for more than a quarter of the study population as well, in which hypertension and diabetes prevailed.

Almost half of the patients (83 [45.9%]) were hospitalized due to conditions unrelated to the SARS-CoV-2 infection which can be explained by patients asymptomatic for COVID-19 having been admitted to hospital for other cancer-related clinical complications. An intra-hospital transmission may also be considered, raising an important issue about the remarkable risk of infection to patients admitted for elective procedures. As for the symptoms present at diagnosis, similarly to data of other series^7,8,10,12,14^, in the current cohort, dyspnea, cough and fever were all highly frequent.

The odds of some COVID-related complications were quite similar to the findings reported by Kuderer et al.^14^ in an international prospective series in which more than 928 patients were analyzed. The rate of ICU admission of 14% (*versus* 17.7% in the current study) and the mechanical ventilation requirement rate of 12% (*versus* 19.3% in the current study) were also alike in proportional terms. Zhang et al.^6^ also showed paralleled data with respect to other variables such as the demand of supplemental oxygen in 78.6% of cases (*versus* 71.8% in the current study).

Early data from non-cancer patients suggested that 14-19% of cases progress with severe complications, such as septic shock, respiratory failure, acute kidney injury and multiple organ failure^5,15,18^. In the present study, these rates were much higher, ranging from 18.2 to 38.7%, highlighting the increased likelihood of severe complications in cancer patients. Conversely, cerebrovascular and cardiovascular events were less frequent.

In total, 69 (38.1%) of 181 patients died. Herein, COVID-19-related mortality was considered as the endpoint. Consequently, nine patients who clearly have recovered from COVID-19, and died due to other cancer-related reasons, were excluded from this mortality analysis. Therefore, the overall COVID-19-related mortality rate reached almost one third of the cases (60 [33.1%]), which was higher than that reported by other series with cancer patients^7,8,10,12,14^, and far exceeding the mortality reported for non-cancer patients^5^. It is important to point out that some patients had the definition of non-invasive support after or even before the diagnosis of COVID-19 due to the severity of their advanced malignancies, which may have overestimated the mortality rate. In addition to this, a noticeable number of the analyzed patients (103 [56.9%]) were under palliative or best supportive care at the time, reflecting the advanced stage of their respective diseases.

Unlike in the multicenter studies recently published by Mehta et al.^10^ and Dai et al.^15^, the mortality rate was higher in patients with solid tumors than in patients with hematological malignancies, possibly due to the small number of hematological patients in this cohort. But likewise, lung cancer, breast cancer and gastrointestinal cancer stood for the highest numbers of cases progressing to death.

In line with the results published by Kuderer et al.^14^, the characteristics associated with clinical fragility, such as being elderly, having advanced stage, a greater number of metastases, pulmonary metastases, palliative or supportive treatment and symptomatic patients at hospital admission were significantly associated with a higher risk of death. In this same context, the type of anti-cancer treatment received by patients within the previous 60 days did not influence survival outcome.

Except for the association of C-reactive protein with mortality (OR 1.04; p = 0.002), none of the laboratory markers were likely to predict a higher risk of mortality. Other laboratory exams, including inflammatory markers such as lactate, ferritin, fibrinogen, and lactate dehydrogenase were not regularly collected in this cohort, preventing related analyses. A prospective study in order to evaluate the immune response markers in our cohort of cancer patients with COVID-19 is being currently conducted.

As in the study performed by Kuderer et al.^14^, none of the specific therapies prescribed such as antiviral oseltamivir (used for the initial suspicion of influenza infection), therapeutic anticoagulation, ivermectin or chloroquine influenced the risk of death in the current cohort. The strong association between the use of antibiotics and the outcome of death can be explained by the fact that these patients showed a more serious condition than COVID-19, including coinfections.

Some important limitations are also worth mentioning in this study. As a single-center cohort in a country of continental proportions, such as Brazil, a selection bias may well exist, hindering an external validity. The missing data rate for some variables was considerably high due to the retrospective design of the study. There was no paired sample with non-cancer patients with COVID-19 or cancer patients without COVID-19 to provide a better comparison between the outcomes of morbidity and mortality. Due to the in-hospital follow-up only, there was no report of long-term morbidity. Finally, the general population of the study was very heterogeneous with several types of neoplasia and anti-cancer treatment, making it difficult to design a more reliable portrait by tumor site.

Lastly, finishing on a positive note, some strengths of the current study can also be recognized. The Brazilian National Cancer Institute is the most important national reference center for the treatment of cancer patients through the Brazilian Public Health System (SUS), with a high admission charge, enabling a quick inclusion of patients for this study. This was one of the largest series ever undertaken to explore the impacts of SARS-CoV-2 infection specifically in cancer patients. Throughout the analysis, many variables were presented, allowing us to explore the possibility of their association with risk of death. Ultimately, this is the first set of Brazilian data in this field, ever.

Conclusively, like other comorbidities, cancer is suggested to be an important prognostic factor for patients with COVID-19, probably due to the greater clinical fragility and the negative impact of immunosuppressive treatments. Despite having formulated early institutional emergency measures, since the onset of the pandemic in Brazil, to reduce the exposure of this group of patients to SARS-CoV-2, INCA struggled with the considerable burden of infected patients in need of hospitalization. There is an urgent demand for an ostensible national strategy in order to lower the spread of virus among cancer patients, prioritizing social isolation, surveillance and mass testing for SARS-CoV-2.

## Data Availability

All data will be made available upon written request to the Corresponding author by e-mail.

## Contributors

The study design was planned by ACM, LCST, MAS and JPBV; ACM, JLS and LZA were involved in the data collection; ACM, LCST and JLS in analysis and interpretation. All authors (ACM, LCST, JLS, LZA, ACPS, LORR, MSC, MMG, GLQM, ACPMP, MAS and JPBV) contributed to the writing, critical reviewing and submission of the manuscript.

## Data sharing

Our data are accessible to researchers upon request for data sharing to the corresponding author.

## Acknowledgments

The authors thank all the INCA health workers for taking care of cancer patients with COVID-19. This work was supported by grants to LCST from CNPq (306798/2019-0), to MAS from CNPq (305765/2015-9) and FAPERJ (E-26/202.894/2017) and to JPBV from CNPq (307042/2017-0) and FAPERJ (202.640/2019, 211.562/2019 and 010.000162/2020). LZA was supported by a PIBIC/CNPq fellowship.

